# Estimation of SARS-CoV-2 Infection Prevalence in Santa Clara County

**DOI:** 10.1101/2020.03.24.20043067

**Authors:** Steve Yadlowsky, Nigam Shah, Jacob Steinhardt

## Abstract

To reliably estimate the demand on regional health systems and perform public health planning, it is necessary to have a good estimate of the prevalence of infection with SARS-CoV-2 (the virus that causes COVID-19) in the population. In the absence of wide-spread testing, we provide one approach to infer prevalence based on the assumption that the fraction of true infections needing hospitalization is fixed and that all hospitalized cases of COVID-19 in Santa Clara are identified.

Our goal is to estimate the prevalence of SARS-CoV-2 infections, i.e. the true number of people currently infected with the virus, divided by the total population size.

Our analysis suggests that as of **March 17, 2020**, there are **6**,**500 infections (0.34% of the population)** of SARS-CoV-2 in Santa Clara County. Based on adjusting the parameters of our model to be optimistic (respectively pessimistic), the number of infections would be 1,400 (resp. 26,000), corresponding to a prevalence of 0.08% (resp. 1.36%). If the shelter-in-place led to R_0_ < 1, we would expect the number of infections to remain about constant for the next few weeks. However, even if this were true, we expect to continue to see an increase in hospitalized cases of COVID-19 in the short term due to the fact that infection of SARS-CoV-2 on March 17th can lead to hospitalizations up to 14 days later.

## Introduction

Inference of the prevalence of SARS-CoV-2 in the US is complicated by the lack of widespread testing. Testing is unreliable for providing prevalence of the disease in the entire population, because tested individuals are not representative of the population at large. Tested individuals are selected based on symptoms, and are necessarily a subset of the total number of individuals infected. There may be many individuals with few to no symptoms that do not get tested, but nevertheless are vectors for the SARS-Cov-2 virus.

Testing in the Bay Area has been reliable enough that most individuals hospitalized for pneumonia or other complications caused by COVID-19 are likely to be tested, and positively identified. Therefore, we can use these data, along with the hospitalization rate of COVID-19 estimated from other countries to infer the number of cases in the area that would lead to this level of hospitalization.

Also, given the hospitalization data, we can estimate the rate of growth of cases, and project this forward to estimate future hospitalizations. Given the shelter-in-place order for our area, our hope is that R_0_ < 1, starting on March 17th. If we make the optimistic assumption that there are essentially no new infections after the shelter-in-place order, we still expect hospitalizations to increase for 10 to 14 days, leading to a peak hospital bed demand 3x to 16x greater than at the time of the order; our best guess is 6x, but the precise ratio would require modeling time lag from infection to symptoms to hospitalization more precisely, where we defer to ongoing modeling efforts.

## Modeling details

Our model is to assume that the number of hospitalizations at any point in time is proportional to the number of total infections some number of days before, based on the *lag time* between infection and hospitalization.

### Simple model

Therefore, to estimate the number of infections on day *t*, we use the number of hospitalizations *h(t)*, and use the formula *infections(t) = exp(lag time * exponential growth rate) * h(t) / hospitalization rate*. This can be converted to a prevalence fraction by dividing by the population size. Note that the hospitalization rate is needed to estimate the total number of infections, but not for forecasting overall hospital bed demand.

### Incremental model

One might be concerned with the above approach because not all past infections would lead to a hospitalization in lag-time days; only the new infections lead to new admissions. Therefore, we should look at the increments in the number of hospitalizations to calculate the number of new infections from the *lag-time* days prior. Then, we can sum up the number of new infections up to a given date to get the cumulative number of infections. It turns out that because of the linearity of the conversion from hospitalizations to infections, these two approaches will give approximately the same answer. Therefore, we will stick to the simpler model for the results presented here.

### Parameters and data sources

As input parameters to our model, we need an estimate of the *lag time*, and the *rate of growth of infections*, and *hospitalization rate* for COVID-19 among those infected. As input data, we need the *number of hospitalizations*, and the *size of the population* from which those hospitalizations are drawn.

For the **lag time**, we need to combine the incubation time and the time for disease progression to severe symptoms. The median incubation time is estimated to be about 5 days^1^. The time from having symptoms to needing hospitalization is about 1 week, adding up to 12 days. In the Chinese data, the lag between the maximum onset proportion at January 23 to the maximum hospitalization at February 4 is 12 days, matching this estimate^2^. We believe these may be slightly overestimated, and use 11 days in our model.

For the **rate of growth of infections**, we compared two values: the first estimated from the change in hospitalizations from March 3 to March 12 in the Santa Clara data, and the second calculated from the reported 6-9 day doubling time^3,4^. The estimate of the rate of growth of infections from hospitalizations gives a 14.4% growth per day and the estimate from Chinese data gives 8-12% growth per day.

Because increases in the number of tests performed (which is growing quickly at about 30% per day^5^) affects the number of confirmed cases, relying on the growth of confirmed cases in the Santa Clara County will likely overestimate the growth rate of infections, so we use this as our upper bound.

We can approach estimating the **hospitalization rate** in three ways. The first is to use the Imperial College report, which puts hospitalization at about 4.4% ^6^. The second is to use our institution’s hospitalization rate among those who test positive for COVID-19, and adjust for the fact that for many, the disease is mild enough that they do not seek healthcare. If 80% of cases are mild, we can take the Stanford test-hospitalization rate, which is 18.3% (95% CI 11.7%, 24.9%) and divide by 5 to get the COVID-19 hospitalization rate of 3.66%. The third approach is to use the hospitalization rate from China^7^, and adjust for the fact that many infections could have been missed. This value is likely an overestimate due to substantial under-reporting in China ^8 9^.

The **number of hospitalizations** are drawn from Santa Clara county’s reports on the number of hospitalizations, using the Internet Archive [Wayback Machine](http://web.archive.org/). Therefore, these are drawn from a population of 1.938 million people. This population could be larger, or smaller, depending on whether everyone in the county goes to hospitals in the county, and whether these are the only people going to Santa Clara hospitals.

### Parameter uncertainty

Because we do not know many of the parameters exactly, we bracket them between a lower bound and upper bound, and a best guess based on what we know so far. We use each of these (lower bound, upper bound and best guess) to obtain the number of inferred infections and prevalence.

The parameters are summarized below in Table 1. Our lower bounds are a 10.5% increase in infections per day, a 5 day lag time and a 6.2% hospitalization rate of the infected population. Our best guesses are a 15% increase in infections per day, an 11 day lag time, and a 4% hospitalization rate. Our upper bounds are a 22% increase in infections per day, a 12 day lag time, and a 2.3% hospitalization rate.

**Table 1.** Parameters of our model, and our optimistic and pessimistic bounds. Note that because a lower hospitalization proportion leads to a higher estimate of the number of SARS-CoV-2 infections, it is listed in the “Upper Bound Parameters” column.

|  | Lower Bound Parameters | Best Guess Parameters | Upper Bound Parameters |
| --- | --- | --- | --- |
| **Daily case increase** | 10.52% | 15.00% | 22.14% |
| **Infection to Hospitalization Lag** | 5 | 11 | 12 |
| **Hospitalization Proportion** | 0.0624 | 0.0400 | 0.0234 |
| **Population Size** | 1,938,000 | 1,938,000 | 1,938,000 |
| **Estimated Prevalence** | 0.08% | 0.34% | 1.36% |

We can perform a sensitivity analysis under a variety of sampled estimates of these parameters, drawing uniformly over the range specified above, and re-running the analysis a 1000 times. Below, we report the range and quartiles of these analyses.

## Results

The inferred number of infections for March 17 is 6,500, and the lower and upper bounds are 1,400 and 26,000, respectively. These estimates provide a prevalence of 0.34%, with bounds of 0.08% to 1.36% (Table 1). If the shelter-in-place order worked, this would be the expected maximum prevalence in the area, until people recover. Unfortunately, we will not know until about March 27-31 if this is the case, at which point we expect the number of hospitalizations to plateau. The detailed results are in Table 2.

**Table 2.** Hospitalizations reported in Santa Clara County and our estimates of the number of SARS-CoV-2 infected individuals in the county using our model and the parameters reported in Table 1.

| Date | Hospitalized | Imputed Hospitalized Cases | Lower Bound Estimated Infections | Best Guess Estimated Infections | Upper Bound Estimated Infections | Lower Bound Prevalence | Best Guess Prevalence | Upper Bound Prevalence |
| --- | --- | --- | --- | --- | --- | --- | --- | --- |
| 3/2/2020 | 4 | 4 | 106 | 465 | 1885 | 0.01% | 0.02% | 0.10% |
| 3/3/2020 | 11 | 11 | 291 | 1279 | 5184 | 0.02% | 0.07% | 0.27% |
| 3/4/2020 |  | 13 | 333 | 1464 | 5931 | 0.02% | 0.08% | 0.31% |
| 3/5/2020 |  | 14 | 381 | 1675 | 6787 | 0.02% | 0.09% | 0.35% |
| 3/6/2020 |  | 16 | 436 | 1917 | 7767 | 0.02% | 0.10% | 0.40% |
| 3/7/2020 |  | 19 | 499 | 2194 | 8887 | 0.03% | 0.11% | 0.46% |
| 3/8/2020 |  | 22 | 571 | 2510 | 10169 | 0.03% | 0.13% | 0.52% |
| 3/9/2020 |  | 25 | 653 | 2872 | 11637 | 0.03% | 0.15% | 0.60% |
| 3/10/2020 |  | 28 | 747 | 3287 | 13316 | 0.04% | 0.17% | 0.69% |
| 3/11/2020 |  | 32 | 855 | 3761 | 15237 | 0.04% | 0.19% | 0.79% |
| 3/12/2020 | 37 | 37 | 978 | 4303 | 17435 | 0.05% | 0.22% | 0.90% |
| 3/13/2020 | 38 | 38 | 1005 | 4420 | 17907 | 0.05% | 0.23% | 0.92% |
| 3/14/2020 | 48 | 48 | 1269 | 5583 | 22619 | 0.07% | 0.29% | 1.17% |
| 3/15/2020 | 52 | 52 | 1375 | 6048 | 24504 | 0.07% | 0.31% | 1.26% |
| 3/16/2020 |  | 54 | 1428 | 6281 | 25446 | 0.07% | 0.32% | 1.31% |
| 3/17/2020 | 56 | 56 | 1481 | 6513 | 26389 | 0.08% | 0.34% | 1.36% |

Table 3 contains the sensitivity analysis where we consider other combinations of the parameters in the ranges provided, and rerun our analysis with 1000 randomly selected combinations. The results are similar to those reported above, although they cluster closer to the best guess of parameters.

**Table 3.** The estimated number of SARS-CoV-2 infected individuals on March 17 from a sensitivity analysis where we randomly sample parameters from within our range and compute the estimated number of infections.

| Percentile | Estimated Infections | Estimated Prevalence |
| --- | --- | --- |
| 1st | 1634 | 0.08% |
| 25th | 3461 | 0.18% |
| 50th | 4786 | 0.25% |
| 75th | 6781 | 0.35% |
| 99th | 20263 | 1.05% |

## Discussion

It’s unclear if hospitalizations are a reliable source of data or not. One thing we noticed is that the numbers are small, and so directly fitting a model for log(hospitalizations) as a function of days since the outbreak does not give a very good estimate of the growth rate over time. This may be because doing so is sensitive to noise. However, we think that while this may be more sensitive to noise, it is less sensitive to selection biases, and therefore may serve as a more reliable estimate of prevalence than positive testing rates.

There has been some discussion of deploying a randomized testing program. If the prevalence of COVID-19 is between 0.13% and 1.36%, then such a prevalence estimate has implications for the size of the testing necessary to get a reliable estimate of prevalence. To have at least 10 positive samples, somewhere between 1,000 and 10,000 randomly selected individuals would need to be tested. To have enough to have good statistical resolution may require many more. However, doing at least 5,000 tests would help us to identify if we are in the right ballpark, in terms of prevalence.

One question that comes up from these analyses is when would we expect the number of infections to plateau after the shelter-in-place order, if the order were to stop or reduce the spread to below exponential growth. Such an order should immediately affect the number of infections that we projected here, so that the number of infections does not grow beyond that of March 17th, and starts to dwindle after the 14-21 day course of the virus infection. However, because of the lag between infection and hospitalization, we expect the number of new hospitalizations to continue to increase for another 12 days. The lag time varies between 3 and 12 days^1^, therefore, we would expect to see a slight change in rate of increase of hospitalizations in about 1 week from March 17th; the number of hospitalizations will still continue to increase and only the rate of increase will slow. Due to such large variance in the number of days at which people present at the hospital, reading too much into the day to day numbers and reactively changing policy before the 12 days after the shelter-in-place may not be appropriate.

## Data Availability

All publicly available data included in results.

## Acknowledgements

We thank Robert Tibshirani for his help with running the sensitivity analysis, and providing useful comments on this report. We also thank various people in the Stanford Medicine community for their help curating the reports and papers that we cite for selecting the parameters in our model.

